# Galectin-3 and Heart Failure with Preserved Ejection Fraction: Clarifying an Emerging Relationship

**DOI:** 10.1101/2023.01.11.23284456

**Authors:** Basil M. Baccouche, Emmajane Rhodenhiser

**Author notes:** **Corresponding Author** Emmajane Rhodenhiser, BS Neurosurgery Clinical Research Assistant, 185 Meeting St, Providence, RI, 02912.

## Abstract

**Introduction:** HFpEF is one of the leading causes of death whose burden is estimated to expand in the coming decades. This paper examines the relationship between circulating levels of galectin-3, an emerging risk factor for cardiovascular disease, and the clinical diagnosis of HFpEF.

**Methods:** The authors reviewed peer-reviewed literature and 18 studies met inclusion criteria. Study characteristics, study outcome definitions, assay characteristics, main findings, and measures of association were tabulated and summarized.

**Results:** Five (1–5) studies found significant associations between galectin-3 and HFpEF diagnosis compared to healthy controls, and one (6) did not. Five studies (7–11) found significant associations between galectin-3 concentration in circulation and severity of diastolic dysfunction. Three studies (12–14) found a statistically significant association between circulating galectin-3 and all-cause mortality or rehospitalization. Two studies (15,16) found levels of circulating galectin-3 to be a statistically significant predictor of later HFpEF onset. Finally, two studies examined whether galectin-3 was associated with incident HFpEF, one (17) found a significant association and the other (18) did not.

**Conclusion:** Given the paucity of effective therapeutics for HFpEF, galectin-3 shows promise as a possible HFpEF-linked biomarker that may, with further study, inform and predict treatment course to reduce morbidity and mortality.

## INTRODUCTION

Cardiovascular disease (CVD) is the number-one cause of death in the United States (19– 22). Heart failure with preserved ejection fraction (HFpEF) is a type of CVD in which heart failure symptoms are observed but the fraction of blood ejected to systemic circulation is conserved, likely due to impaired ventricular filling (23–25). The burden of HFpEF has increased in the last decade, currently representing a majority of all cases of HF (26,27). The global burden is anticipated to increase as well in the coming decades (28). Despite this alarming trend, there are still no effective therapies for HfpEF (29).

Galectin-3, classified in the galectin family, is a protein that has been shown to be causally linked to pathophysiological cardiovascular processes, including atherosclerosis, fibrosis, and heart failure (30). Galectin-3 has been presented in the literature as a novel biomarker for cardiac disease diagnosis, and a recent meta-analysis has shown that levels of circulating galectin-3 are associated with incident heart failure (31,32).

As the global burden of heart disease continues to grow, so too does the importance of advancing preventative tools like biomarker-based risk prediction models. An incomplete understanding of the molecular and pathophysiological pathways underlying the development of HFpEF may be one of many factors limiting the development of an effective HFpEF therapy.

The aim of this article is to interrogate and clarify the relationship between HFpEF diagnosis and concentration of circulating galectin-3 through a rigorous, reproducible review of the published literature associating HFpEF-related-endpoints and galectin-3. The authors, to the best of their knowledge, have not identified any review examining this specific relationship and thus hope that this synthesis of findings will prove useful for the investigator interested in galectin-3 as a potential biomarker for HFpEF.

## METHODS

The Sciome Workbench for Interactive computer-Facilitated Text-mining (SWIFT)-Review, was used to perform a review examining the association between levels of galectin-3, a circulating biomarker, and incidence of heart failure with preserved ejection fraction (HFpEF) (30,33,34). SWIFT is an efficient tool which uses statistical text mining to make it easier to manually screen results. Table 1 outlines the search terms that were processed within the National Library of Medicine’s MEDLINE database: [(galectin-3 OR gal-3) AND (HFpEF)], with zero restriction settings applied to the search tool. Because of the low number of total results (45), automated screening results were double checked via manual screening by the authors. Study inclusion criteria are shown in Table 1, and a PRISMA-compatible flow chart is included below in Figure 1 for transparency and reproducibility.

**Table 1:** Study Inclusion Criteria.

| Inclusion Criteria |
| --- |
| • Human subject research<br>• English language<br>• Full-text freely available to University of Cambridge<br>• HFpEF clearly defined and diagnosed<br>• Includes the exposure galectin-3<br>• Includes the outcome HFpEF diagnosis<br>• Primary research |

**Figure 1:**
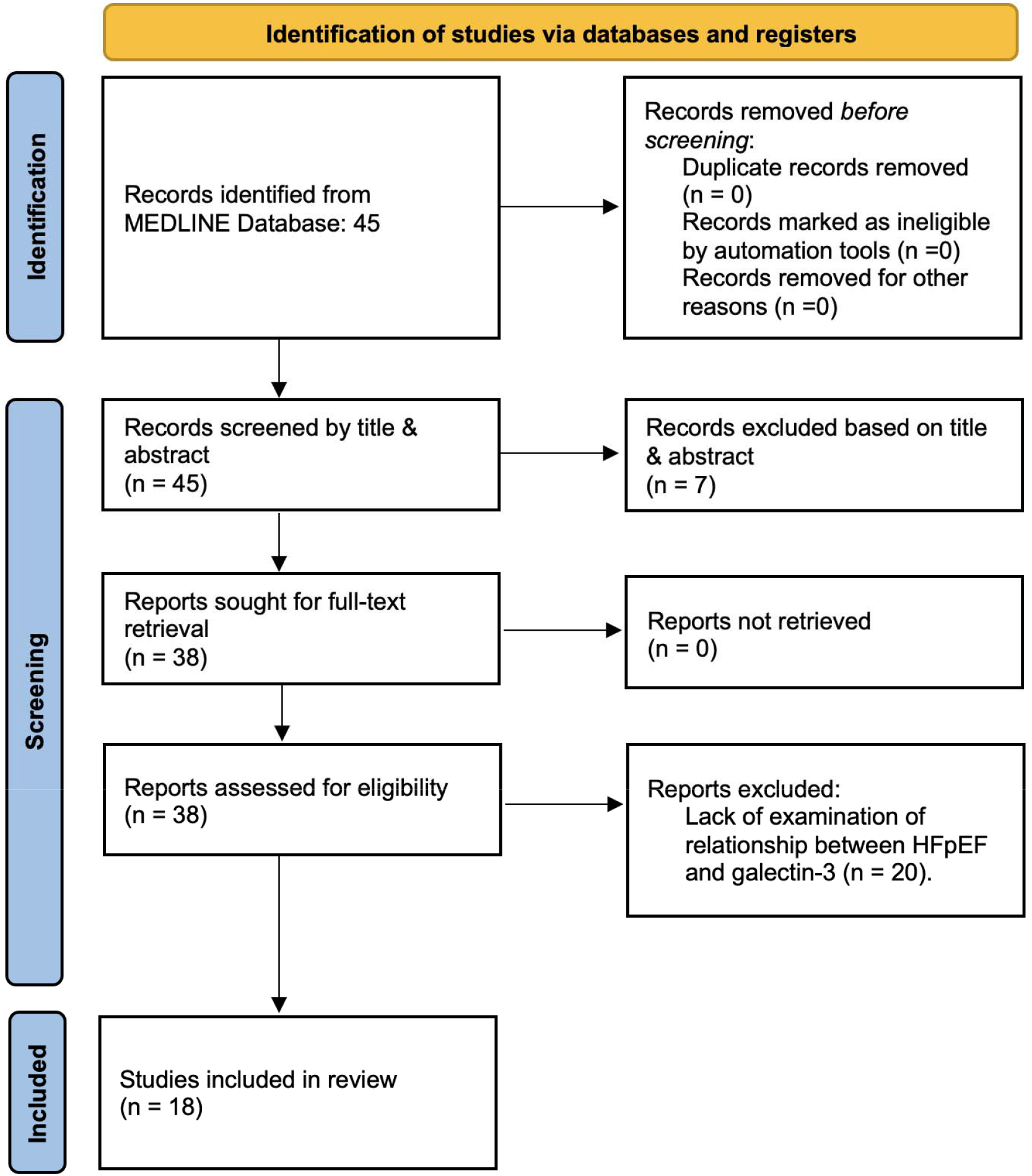
Reproducible, PRISMA-compatible review workflow(35). All intra-study data were extracted manually by the authors.

## RESULTS

### Characteristics

18 studies met the inclusion criteria outlined in Table 2. Amongst the 18 qualifying studies, six were case-control studies, seven were prospective cohort studies, four were cross-sectional, and one was a retrospective cohort study. Sample size across all 18 studies ranged from 62 and 22,756, and the average participant age was between 55.9 and 75 years. 12 of the 18 study populations included diabetics. Studies were conducted in the China, Europe, India, Russia Taiwan and United States. Table 2 summarizes study.

**Table 2:** Study characteristics.

| **Authors** | **Study Design** | **Study Period** | **Geographic Location(s)** | **Population Source(s)** | **Sample Size** | **Mean Age** | **% Female** | **% Diabetics** |
| --- | --- | --- | --- | --- | --- | --- | --- | --- |
| Yin et al., 2014(3) | Case Control | 2013 | China | Community-based | 78 | 71.86 | 11.50% | 42.31% |
| Wu et al., 2015(8) | Cross-Sectional | N/A | Taiwan | Community-based | 176 | 68.23 | 38.07% | N/A |
| Yu et al., 2015(13) | Prospective Cohort | 2010-2011 | China | Community-based | 261 | 70.02 | 50.96% | N/A |
| Edelmann et al., 2015(14) | Prospective Cohort | 2007-2011 | Germany & Austria | Community-based (Aldo-DHF) | 415 | 67.00 | 52.30% | 16.60% |
| Berezin et al., 2016(16) | Case Control | 2012-2015 | Ukraine | Community-based | 199 | 55.90 | 47.70% | 19.10% |
| Beltrami et al., 2016(9) | Prospective Cohort | 2012-2015 | Italy | Community-based | 98 | 74.84 | 51.02% | 34.69% |
| Polat et al., 2016(4) | Case Control | 2013-2014 | Turkey | Community-based | 82 | 58.61 | 46.34% | 34.15% |
| de Boer et al., 2018(17) | Prospective Cohort | CHS: 1989-1990,1992-1993<br>FHS:1995-1998<br>MESA:2000-2002<br>PREVEND:1997-1998 | United States | Community-based (FHS, CHS, PREVEND, and MESA) | 22756 | 60.00 | 53.12% | 10.00% |
| Wu et al., 2018(10) | Cross Sectional | 2011-2015 | Taiwan | Community-based | 77 | 67.79 | 55.86% | N/A |
| Cui et al., 2018(12) | Case Control | 2014-2016 | China | Community-based | 247 | 71.93 | 54.70% | 30.77% |
| Ansari et al., 2018(7) | Prospective Cohort | 2014-2016 | Germany | Community-based | 70 | 65.00 | 49.00% | 24.00% |
| Lebedev et al., 2020(5) | Case Control | N/A | Russia | Community-based | 62 | 59.29 | 43.55% | 100.00% |
| Merino-Merino et al., 2020(6) | Cross Sectional | 2015 - 2017 | Spain | Community-based | 115 | 62.78 | 30.43% | 13.91% |
| Pecherina et al.,<br>2020(11) | Prospective<br>Cohort | 2015 | Russia | Community-based | 254 | 60.64 | 30.71% | 16.14% |
| Mitic et al.,<br>2020(2) | Cross<br>Sectional | 2018 | Serbia | Community-based | 112 | N/A | N/A | N/A |
| Kanukurti et al.,<br>2020(1) | Case Control | N/A | India | Community-based | 83 | 57.25 | N/A | N/A |
| Watson et al.,<br>2021(15) | Retrospective<br>Cohort | 2009-2011 | Ireland | Community-based<br>(STOP-HF) | 90 | 75.00 | 53.30% | N/A |
| Trippel et al.,<br>2021(18) | Prospective<br>Cohort | 2004-2016 | Germany | Community-based | 1386 | 67.00 | 50.90% | 26.60% |

### Outcomes

All 18 studies included clinical diagnosis of HFpEF as a variable of interest. Outcomes were ascertained using validated standardized criteria, except for two studies (1,6) which used HFpEF as diagnosed internally by cardiologist(s). All studies used echocardiographic data in combination with other clinical measurements to confirm diagnosis of HFpEF. Study-specific HFpEF definitions and outcome ascertainment methods are shown in Table 3.

**Table 3:** Study outcomes.

| Authors | Defining Outcome of Interest (HFpEF) | Follow-Up Length in Years | Outcome Collection | Outcome Ascertainment |
| --- | --- | --- | --- | --- |
| Yin et al., 2014 | (1) LVEF > 45% (2) standard size of the left ventricular cavity, and (3) left ventricular diastolic dysfunction as confirmed by echocardiography. Functional classification per NYHA. | N/A | Echocardiography, medical records, plasma samples. | NYHA |
| Wu et al., 2015 | HF as previously diagnosed site. HFpEF severity as determined by Doppler imaging (E/Em) and CMRI. | N/A | Cardiac magnetic resonance imaging, doppler imaging, echocardiography, hemodynamic data, medical records, plasma samples. | Framingham criteria |
| Yu et al., 2015 | HF as diagnosed per ESC 2008, and functional classification as assigned according to NYHA. | 12 months | Angiography, clinical assessment, electrocardiography, telephone interviews. | ESC, NYHA |
| Edelmann et al., 2015 | HFpEF as previously defined by the Aldo-DHF study. There were four criteria for participation: (1) NYHA class II or III (2) LVEF ≥ 50%, (3) Grade ≥ I diastolic dysfunction 22 or current AF, per echocardiogram and (4) peak VO<sub>2</sub> ≤ 25 mL/kg/min. | 12 months | Echocardiography, medical records, plasma samples. | NYHA |
| Berezin et al., 2016 | Chronic HF as per contemporary clinical criteria. HFrEF (LVEF ≤ 45%) and HFpEF (LVEF ≥ 50%). | N/A | Echocardiography, doppler Imaging, medical records, serum samples. | ESC |
| Beltrami et al., 2016 | All patients had (1) new onset or exacerbated AHF, (2) acute decompensated HF per signs and symptoms. HFrHF (EF below 50%) or HFpEF (EF above 50%) per echocardiogram. | 6 months | Anamnestic investigation, clinical assessment, echocardiography, plasma samples, telephone calls. | ASE |
| Polat et al., 2016 | HFpEF as defined by (1) history of symptoms correspondant to NYHA class II or III, (2) LVEF > 50% and LVEDVI < 97 ml/m<sup>2</sup>, (3) LV diastolic dysfunction per echocardiogram | N/A | Clinical assessment, echocardiography, serum samples. | ESC, NYHA |
| de Boer et al., 2018 | Heart failure as determined through signs and symptoms. Incident HF event as categorized by HFpEF (LVEF≥50%), HFrEF (LVEF, <50%), or unclassified. | 144 months | Clinical assessment, echocardiography, medical records, plasma samples. | CHS: CHS criteria, ICD-9 codes 427-429.9. FHS: Framingham criteria. MESA: Identified from records. PREVEND: ESC |
| Wu et al., 2018 | HFpEF as defined by (1) HF defined by Framingham criteria and normal systolic function (ejection fraction ≥ 50%), (2) echocardiographic evidence of LV diastolic dysfunction. | N/A | Cardiac magnetic resonance imaging, doppler imaging, echocardiography, plasma samples. | ESC, Framingham criteria, NYHA |
| Cui et al., 2018 | HF episodes as determined by cardiologists based on biomarkers concentrations and validated according to ESC. | 12 months | Clinical assessment, echocardiography, medical records, plasma and serum samples, telephone interviews. | ESC, NYHA |
| Ansari et al., 2018 | HFpEF as defined by the CIBER study. Grade of diastolic dysfunction per ASE/EACVI Guidelines. | 12 months | Echocardiography, medical records, blood samples, telephone interviews. | ASE/EACVI, NYHA |
| Lebedev et al., 2020 | HFpEF as defined by ESC 2016 | N/A | Echocardiography, medical records, serum samples. | ESC, NYHA |
| Merino-Merino et al., 2020 | HF as diagnosed by a cardiologist. HFmrEF and HFpEF as diagnosed by LVEF between 40-49.9% versus $>50\%$. | N/A | Echocardiography, medical records, serum samples. | Internal Criteria |
| Pecherina et al., 2020 | HFpEF as defined by (1) no clinical signs of HF (Killip class I) and (2) LVEF ≥ 40%. HFrEF as defined by HF signs (Killip class II-IV) and LVEF $<40\%$. | 0.5 months | Echocardiography, medical records, serum samples. | ESC, Killip class |
| Mitic et al., 2020 | HF as defined by (1) symptoms of HF, BNP plasma concentration ($> 35$ pg/mL) and (2) relevant structural heart changes (LV mass index, LVMI $\geq 115$ g/m<sup>2</sup> for males and $\geq 95$ g/m<sup>2</sup> for females or L atrial dilation $\geq 40$ mm) and/or diastolic abnormality (E/A ratio $< 0.75$ or $\geq 1.5$). | N/A | Blood samples, clinical assessment, electrocardiography, echocardiography, medical records. | ESC, NYHA |
| Kanukurti et al., 2020 | HF as defined by clinical symptoms and HFpEF as defined by EF≥ 50% per echocardiogram. | N/A | Echocardiography, medical records, plasma samples. | Internal Criteria |
| Watson et al., 2021 | HFpEF diagnosis as confirmed by (1) new onset HF symptoms (2) LVEF$>$50% and (3) elevated BNP ($>$100 pg/ml), diastolic dysfunction of the LV per Doppler-echocardiogram. | N/A | Clinical assessment, echocardiography, medical records, phlebotomy, serum samples. | ESC, NYHA |
| Trippel et al., 2021 | HFpEF as defined by presence of signs and symptoms of (1) HF, ≥2 Framingham criteria for HF, (2) a preserved LVEF $>$ 50%, and (3) echocardiographic findings of LV diastolic dysfunction. The diagnosis was established when left atrial volume index $>$34 mL/m<sup>2</sup>, or LV mass index (LVMI) ≥115 g/ m<sup>2</sup> for men and ≥95 g/ m<sup>2</sup> for women, or E/e' ≥ 13, or mean e' septal and lateral wall $<$9 cm/s. | 120 months | Clinical assessment, echocardiography, medical records, plasma samples. | ESC, Framingham criteria |

Acute Heart Failure (AHF); American Society of Echocardiography (ASE); B-type natriuretic peptide (BNP); Cardiovascular Imaging and Biomarker Analyses (CIBER); Cardiac Magnetic Resonance Imaging (CMRI); European Association of Cardiovascular Imaging (EACVI); European Society of Cardiology (ESC); Heart Failure (HF); Heart Failure with Preserved Ejection Fraction (HFpEF); Heart Failure with Reduced Ejection Fraction (HFrEF); Left Ventricular (LV); Left Ventricular End Diastolic Volume Index (LVEDVI); Left Ventricular Ejection Fraction (LVEF); New York Heart Association (NYHA).

To measure galectin-3 concentration in circulation, three studies (6,7,15) used Abbot Laboratories’ Architect System, one study (12) used a human galectin-3 assay kit, another (2) used a Quantikine USA kit, and the rest all used enzyme-linked immunosorbent assays (ELISAs) from various manufacturers. Detailed galectin-3 measurement information, including storage temperature and storage duration (where provided), is shown in Table 4.

**Table 4:**
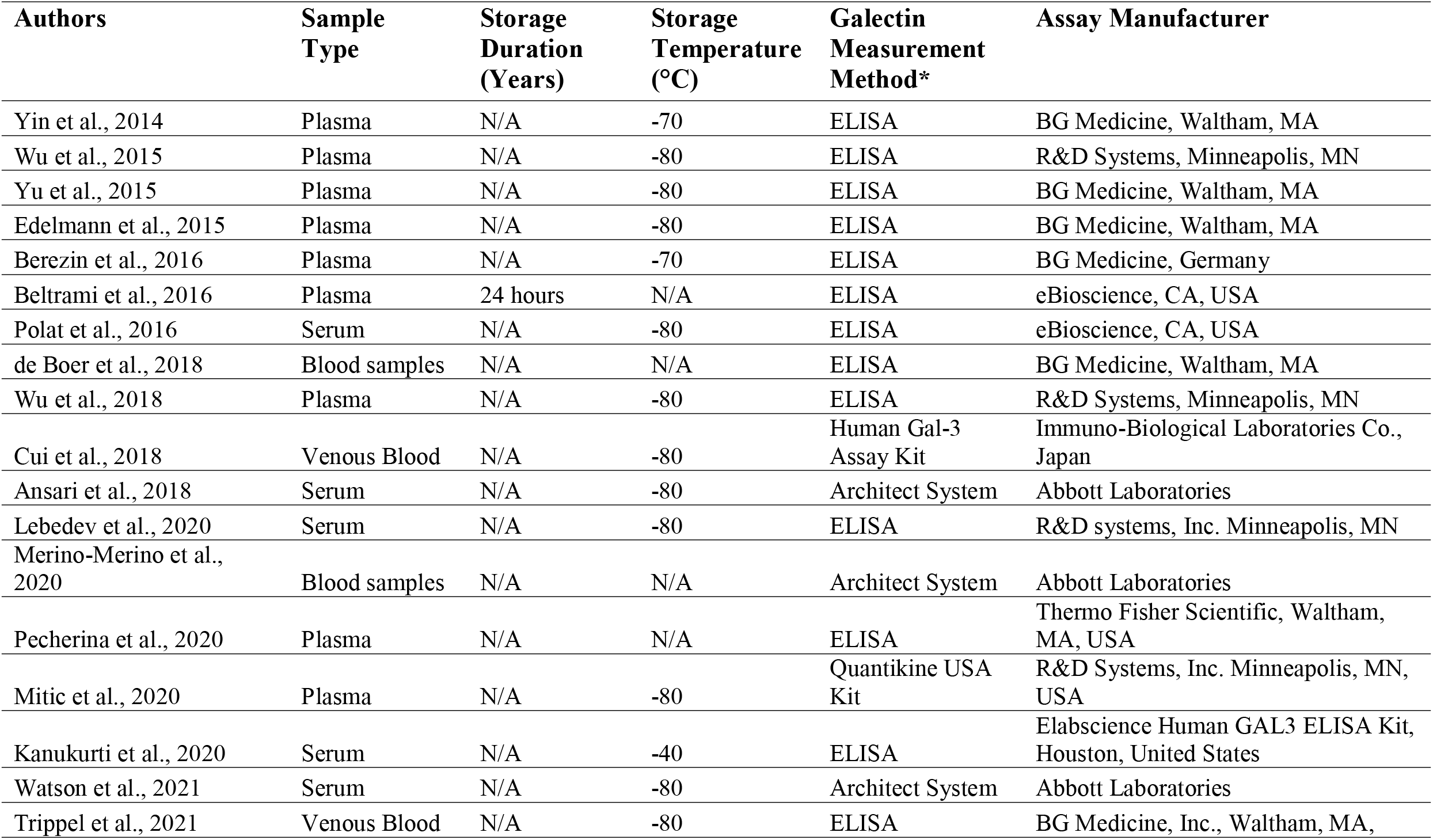
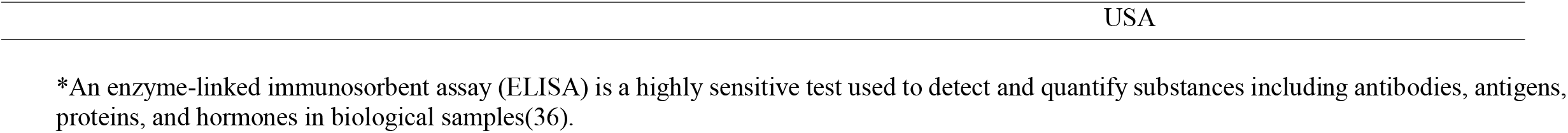
Study assays.

### Main Findings

An overview of main findings from each study is shown in Table 5 and described herewith. Outcome definitions were heterogenous across studies and thus published data were not suitable for meta-analysis. Measures of association (where provided) and p-values, as well as the comparisons under investigation in each study, are shown in Table 6. All studies provided at least one measure of statistical significance (measure of association, p-value, or both).

**Table 5:** Main findings of each study, as reported in the body of the text (and edited for concision).

| Study Authors | Main Findings |
| --- | --- |
| Yin et al., 2014 | Galectin-3 levels were significantly higher in HFpEF patients in comparison to the controls. |
| Wu et al., 2015 | The correlation between plasma and tissue galectin-3 and the degree of diastolic dysfunction and severity of myocardial fibrosis was significant. Cellular galectin-3 responded well to changes in stretching, and tissue galectin-3 responded well to changes in pressure overload. |
| Yu et al., 2015 | Galectin-3 concentration was significantly higher in CHD HF patients. Galectin-3 concentration was an independent predictor of both all-cause mortality and hospital readmittance. |
| Edelmann et al., 2015 | Galectin-3 concentrations are slightly higher in well-compensated HFpEF individuals with predominantly mild symptoms. Galectin-3 concentration is related to different measures of functional performance amongst patients. Galectin-3 levels increase over time in HFpEF patients, which is associated with hospitalization. |
| Berezin et al., 2016 | Galectin-3 plasma concentration is an independent predictor of HFpEF. |
| Beltrami et al., 2016 | Galectin-3 concentrations do not permit distinction between HFrEF and HFpEF individuals. In HFpEF patients, galectin-3 concentration is related to diastolic dysfunction severity and LV stiffness. Independent from renal dysfunction, galectin-3 demonstrates a prognostic role in subjects with HFpEF. |
| Polat et al., 2016 | Galectin-3 was elevated in HFpEF patients in comparison to control patients. Galectin-3 was correlated with NT-proBNP biomarker. Galectin-3 was correlated with L atrial volume index, LV mass index, and E/E'. |
| de Boer et al., 2018 | Galectin-3 concentration was not associated with incident HFpEF. |
| Wu et al., 2018 | Amongst HFpEF patients, galectin-3 was associated with global cardiac fibrosis. |
| Cui et al., 2018 | Galectin-3 level was higher following decreased ejection fraction. Galectin-3 level distinguished HFpEF from controls. Galectin-3 was strongly correlated an increased risk of the endpoint events in HFpEF patients after adjusting for clinical factors and NT-proBNP |
| Ansari et al., 2018 | Galectin-3 corresponded with echocardiogram. Galectin-3 was associated with progressive diastolic dysfunction. Increased galectin-3 levels corresponded to disease course. Galectin-3 levels discriminated patients with grade III diastolic dysfunction on ROC analysis. |
| Lebedev et al., 2020 | Galectin-3 levels were higher in HFpEF and HFmrEF patients as compared to patients without HF. |
| Merino-Merino et al., 2020 | Galectin-3 concentration was not different between HF groups. |
| Pecherina et al., 2020 | Galectin-3 was correlated with diastolic dysfunction parameters in HFpEF patients. Galectin-3 was associated with LVEF and LV end-systolic volume/diameter in HFrEF patients. Galectin-3 levels were higher in HFpEF in comparison to HFrEF patients. |
| Mitic et al., 2020 | Galectin-3 was independently correlated with LV mass index amongst HFpEF patients. Galectin-3 was independently correlated with septum and posterior wall diameters. |
| Kanukurti et al., 2020 | Galectin-3 was higher in HFpEF patients as compared to controls. There was a positive correlation between galectin-3 and NT-ProBNP. Galectin-3 had a 77.78% sensitivity for HFpEF diagnosis. Galectin-3 better than NT-proBNP in diagnosing HFpEF, according to AUC analysis. Galectin-3 was positively correlated with NT-proBNP and lipid parameters. |
| Watson et al., 2021 | Galectin-3 at follow-up was a predictor of incident HFpEF. Galectin-3 over time was a predictor of incident HFpEF. Unadjusted biomarker combinations of galectin-3, hsTroponin-I, and BNP predicted future HFpEF using both baseline and follow-up data. |
| Trippel et al., 2021 | Galectin-3 at baseline was associated with a increased risk for incident HFpEF, cardiovascular hospitalization and death, and overall mortality at ten years of follow-up. |

Area Under the Curve (AUC); B-type natriuretic peptide (BNP); Congenital Heart Defects (CDH); Heart Failure (HF); Heart Failure with Preserved Ejection Fraction (HFpEF); Heart Failure with Reduced Ejection Fraction (HFrEF); High Sensitivity Troponin-I (hsTroponin-I); Left Ventricular (LV); Left Ventricular Ejection Fraction (LVEF); NT-proB-type Natriuretic Peptide (NT-proBNP); Receiver operating characteristic (ROC).

**Table 6:** Measures of association (if provided) of included studies. If only one measure of association was provided, it was considered “further-adjusted”.

| **Study Authors** | **Minimally Adjusted HR [95% CI] (Covariates)** | **Further Adjusted HR [95% CI], P-value** | **Comparison** | **Covariates Used in Further Adjusted Model** |
| --- | --- | --- | --- | --- |
| Yin et al., 2014 | N/A | N/A, p=0.000 | Galectin-3 and HFpEF diagnosis vs controls (no HF) | N/A |
| Wu et al., 2015 | N/A | N/A, p<0.001 | Galectin-3 and severity of diastolic dysfunction | Age, diabetes, gender, LV mass index, plasma NT-proBNP and prescribed drugs |
| Yu et al., 2015 | N/A | *RR: 1.231, 95% CI: 1.066-1.442) | Galectin-3 and all-cause mortality and rehospitalization | N/A |
| Edelmann et al., 2015 | N/A | 3.319 [1.214-9.07] p=0.019 | Galectin-3 and all-cause death or hospitalization | Peak VO<sub>2</sub>, six min walk distance, and short form 36 physical function |
| Berezin et al., 2016 | N/A | 1.08 [1.03–1.12] p=0.002 | Galectin-3 and prediction of HFpEF diagnosis | Diabetes type 2, mellitus, obesity, previous myocardial infarction |
| Beltrami et al., 2016 | HR: 23.98 [3.03–89.45]; p = 0.001 | 19.62 [2.39–60.89] p=0.006 | Galectin-3 and severity of diastolic dysfunction | CKD, diabetes, dyslipidemia, hypertension, smoker |
| Polat et al., 2016 | N/A | N/A, p<0.0001 | Galectin-3 and HFpEF diagnosis vs controls (no HF) | N/A |
| de Boer et al., 2018 | N/A | 1.02 [0.93-1.12] p=0.13 | Galectin-3 and incident HFpEF | Age, BMI, diabetes, hypertension treatment, L ventricular hypertrophy, L bundle branch block, previous myocardial infarction, race/ethnicity, sex, systolic blood pressure, smoking |
| Wu et al., 2018 | N/A | *OR: 1.05 [1.02 - 1.09] p = 0.005 | Galectin-3 and severity of diastolic dysfunction | Diabetes, Endothelin-1, Heart failure, MMP-2, NT-proBNP, TIMP2 |
| Cui et al., 2018 | N/A | 2.33 [1.72–2.94] p=0.009 | Galectin-3 and all-cause death or hospitalization | Age, aldosterone receptor antagonist, b-blockers treatment, coronary artery disease, diastolic blood pressure, eGFR levels, hypertension, LDL cholesterol levels, LVEF, NT-proBNP levels, NYHA grade, sex, systolic blood pressure |
| Ansari et al., 2018 | N/A | *OR: 6.19 [1.489–25.744] p= 0.012 | Galectin-3 and severity of diastolic dysfunction | Age, gender, NT-proBNP, and serum creatinine |
| Lebedev et al., 2020 | N/A | N/A, p=0.01 | Galectin-3 and HFpEF diagnosis vs controls (no HF) | N/A |
| Merino-Merino et al., 2020 | N/A | N/A, p=0.06 | Galectin-3 and HFpEF diagnosis vs controls (no HF) | Age, arterial hypertension, diabetes, obesity, and sex |
| Pecherina et al., 2020 | N/A | N/A, p<0.0001 | Galectin-3 and severity of diastolic dysfunction | N/A |
| Mitic et al., 2020 | N/A | N/A, p<0.001 | Galectin-3 and HFpEF diagnosis vs controls (no HF) | Age, BMI, GDF-15, sST2, and syndecan-1 |
| Kanukurti et al., 2020 | N/A | N/A, p<0.0001 | Galectin-3 and HFpEF diagnosis vs controls (no HF) | Age, comorbidities, sex, and troponin |
| Watson et al., 2021 | 1.15 [1.03-1.28] | *OR: 1.17 [1.02–1.34] p=0.027 | Galectin-3 and prediction of HFpEF diagnosis | Age, sex, levels of: hsTropI, IL6, and ln(BNP) and sST2, |
| Trippel et al., 2021 | N/A | *OR: 1.77 [1.14–2.74] p=0.010 | Galectin-3 and incident HFpEF | Age, BMI, diabetes mellitus, hypertension, kidney function, and sex |

Angiotensin-converting enzyme (ACE); Body Mass Index (BMI); B-type Natriuretic Peptide (BNP); Confidence Interval (CI); Chronic Kidney Disease (CKD); Estimated Glomerular Filtration Rate (eGFR); Growth/Differentiation Factor-15 (GDF-15); Hazard Ratio (HR); Heart Failure (HF); Heart Failure with Midrange Ejection Fraction (HFmrEF); Heart Failure with Preserved Ejection Fraction (HFpEF); High Sensitivity Troponin I (hsTropI); Interleukin-6 (IL6); Left Ventricular (LV); Low Density Lipoprotein (LDL); Matrix Metalloproteinase-2 (MMP-2); New York Heart Association (NYHA); N-terminal-pro hormone B-type Natriuretic Peptide (NT-proBNP); Risk Ratio (RR); Soluble Suppression of Tumorigenicity-2 (sST2); Standard Deviation (SD); Tissue Inhibitor of Metalloproteinases-2 (TIMP2).
\*These studies provided an odds ratio (OR) or risk ratio (RR) as a measure of association instead of a hazard ratio.

Galectin-3 levels were compared with one of five endpoints: levels in healthy controls, severity of diastolic dysfunction, all-cause mortality or rehospitalization, development of HFpEF, and prediction of HFpEF diagnosis. Of the total 18 included studies, six investigated the relationship between levels of circulating galectin-3 and HFpEF diagnosis compared to healthy controls; five(1–5) of those six found statistically significant associations, and one(6) did not. Five studies probed the relationship between galectin-3 concentrations and severity of diastolic dysfunction and all five(7–11) found statistically significant associations. Three studies examined the relationship between levels of circulating galectin-3 and all-cause mortality or rehospitalization, and all three (12–14) found statistically significant associations. Two studies (15,16) found levels of circulating galectin-3 to be a statistically significant predictor of later HFpEF onset. Finally, two studies examined whether levels of circulating galectin-3 were associated with current HFpEF; one (17) found an association that did not meet the threshold of statistical significance, and the other (18) found a significant association.

## DISCUSSION

### Study Limitations

The authors did not review pre-prints journals not indexed in MEDLINE. Galectin-3 is a biomarker associated with a range of cardiovascular dysfunctions and is therefore likely to be limited in utility to use in combination with other diagnostic tools to determine wall thickness, symptomatology, or ejection fraction. The relative lack of studies investigating this association means that the few studies that are available investigate disparate HFpEF endpoints and so are not suitable for pooled meta-analysis, even by HFpEF endpoint subgroup (as sample sizes are small). In addition, each of the 18 included studies is subject to its own methodological limitations, shown in Table 7.

**Table 7:**
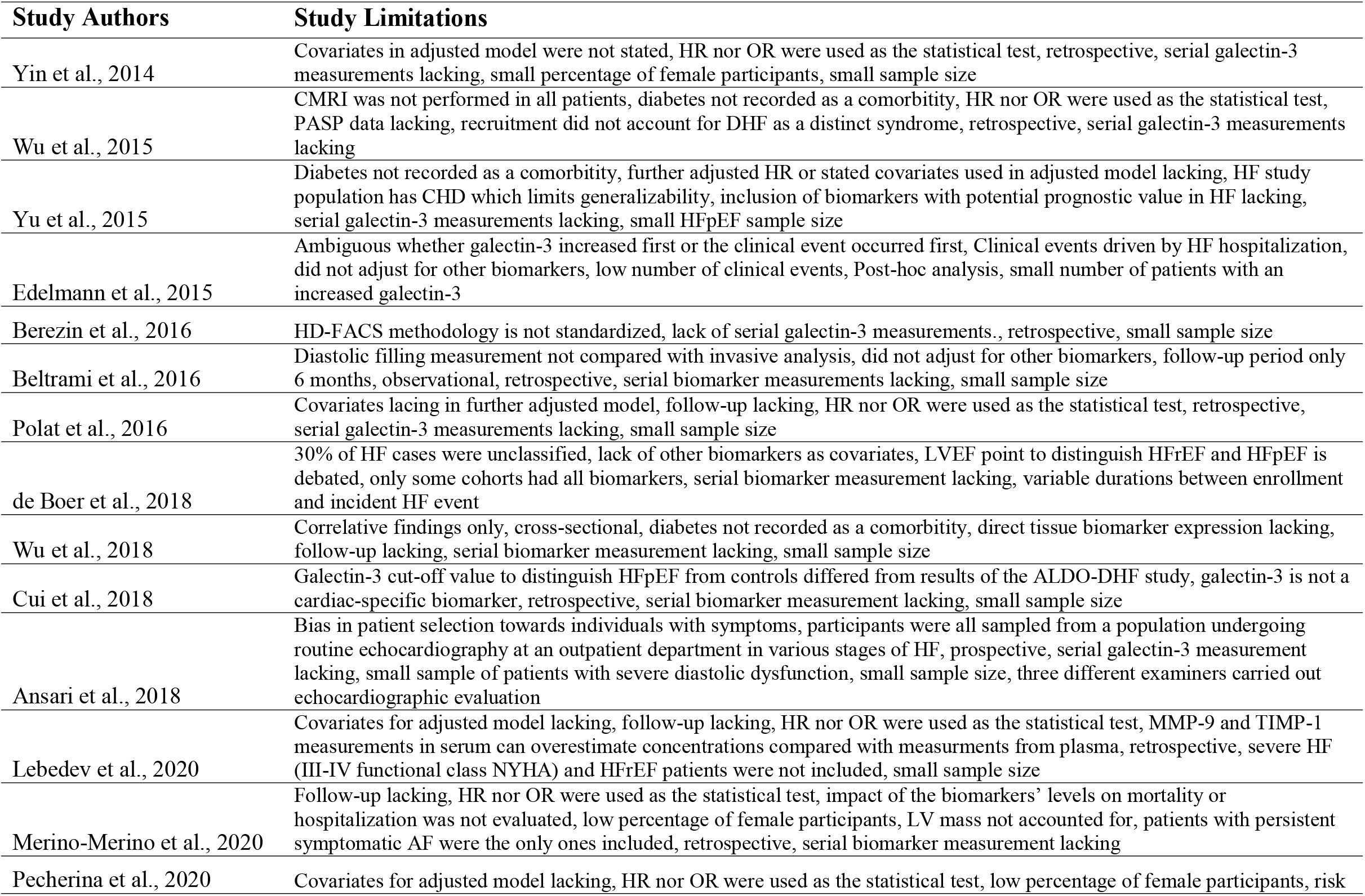
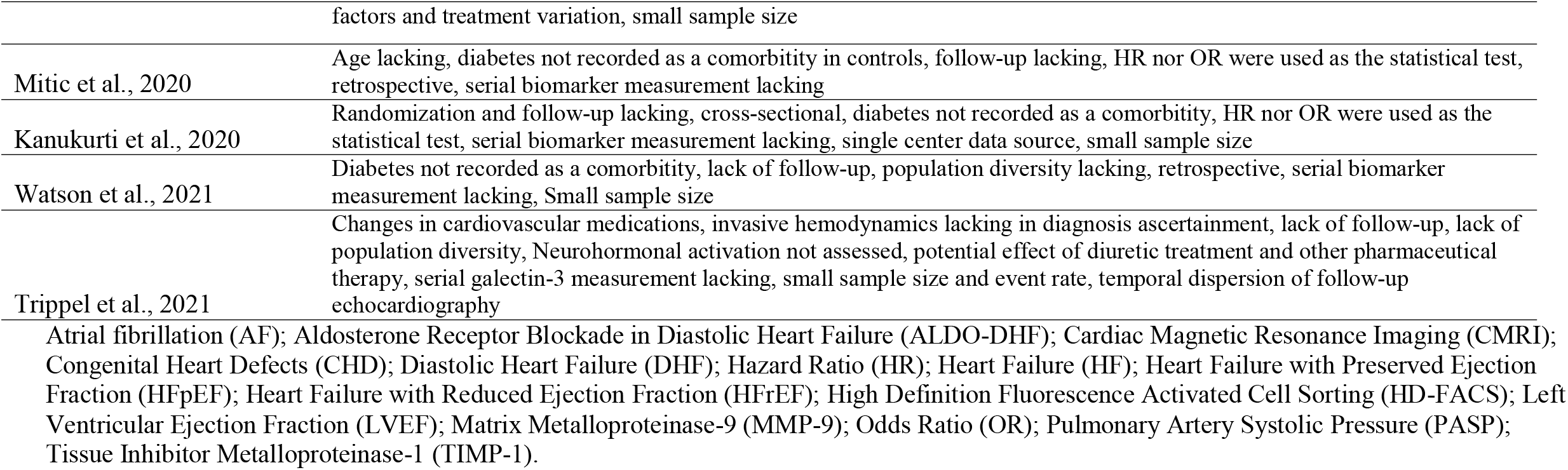
Limitations of included studies, as reported in the body of the text (and edited for concision) and as identified by authors.

### Findings & Implications

This review demonstrates a statistically significant and consistent pattern of association between circulating galectin-3 concentration and respective HFpEF endpoint (incident HFpEF, severity of diastolic dysfunction as assessed by echocardiography or other clinical measurements, or all-cause mortality/rehospitalization due to HFpEF). Only two of the 18 studies found borderline or statistically insignificant associations, and none of the studies reported an inverse association between galectin-3 and HFpEF endpoint. There is an immense, rapidly-growing burden of HFpEF, a well-documented lack of effective treatment, a relative paucity of studies investigating this promising relationship, and a high heterogeneity in HFpEF endpoint across studies which do investigate this relationship (limiting meta-analysis). Therefore, the authors suggest that emerging biomarker galectin-3 – which has been implicated in the pathogenesis of cardiovascular remodeling (31) – should be rigorously interrogated in a large cohort using metrics of risk prediction for HFpEF.

HFpEF is a complex physiological phenomenon and is unlikely to be univariately associated to galectin-3 (or any single biomarker). It is certainly the case that a concert of contributing factors is responsible for the diverse physiological dysfunctions and subsequent symptoms frequently reported in this debilitating disease. However, in seeking to unravel the question of how and when patients develop HFpEF (at which point, it is too late for clinically meaningful reversal of symptoms), the authors propose that this review makes a strong case for galectin-3’s inclusion among suspected biomarkers.

Of the two studies which did not report a statistically significant association, one study (17), while high in sample size, pooled hazard ratios across four studies. One of those four studies (MESA) did not have data for inclusion in this pooled measure of association. Two of the remaining three studies reported a significant association between galectin-3 and incident HFpEF, and the third found no association; when pooled, no association prevailed. Finally, they limited inclusion of HFpEF diagnoses in the pooled dataset only to individuals presenting with HF and undergoing left ventricular function assessment, resulting in 30% of cases with unclassified HF (17). As a result, although this study had an impressive sample size, it is recommended that the relationship between incident HFpEF and galectin-3 in particular be interpreted with caution. The other study which did not report a statistically significant association (6) reported a p-value of 0.06, just above the predefined significance threshold; an argument may be made, given its borderline p-value and modest sample size (n=115), for its potential inclusion within the domain of clinical significance.

### Conclusion

In a review of 18 studies examining relationships between circulating galectin-3 and HFpEF diagnosis, diastolic dysfunction severity, incident HFpEF, or all-cause mortality/rehospitalization, 16 found statistically significant associations and 2 found borderline non-significant associations that are nevertheless of clinical interest. Given the scarcity of effective therapeutics for HFpEF, galectin-3 shows promise as a possible HFpEF-linked biomarker deserving of further study.

## Data Availability

All data in the present work are contained in the manuscript.

## ACKNOWLEDGEMENT

The authors have no financial relationships to disclose.

